# Usability and acceptability of a corneal-plane α-opic light logger in a 24-hour field trial

**DOI:** 10.1101/2023.04.17.23288692

**Authors:** Eljoh Balajadia, Sophie Garcia, Janine Stampfli, Björn Schrader, Carolina Guidolin, Manuel Spitschan

## Abstract

Exposure to light fundamentally influences human physiology and behaviour by synchronising our biological clock to the external light-dark cycle and controlling melatonin production. In addition to well-controlled laboratory studies, more naturalistic approaches to examining these “non-visual” effects of light have been developed in recent years. As naturalistic light exposure is quite unlike well-controlled stimulus conditions in the laboratory, it is critical to measure light exposure in a person-referenced way, the ‘spectral diet’. To this end, light loggers have been developed to capture personalised light exposure. As an alternative to light sensors integrated into wrist-worn actimeters, pendants or brooch-based light loggers, a recently developed wearable light logger laterally attached to spectacle frames enables the measurement of biologically relevant quantities in the corneal plane. Here, we examine the usability and acceptability of using the light logger in an undergraduate student sample (n=18, mean±1SD: 20.1±1.7 yrs; 9 female; Oxford, UK) in real-world conditions during a 24-hour measurement period. We probed the acceptability of the light logger using rating questionnaires and open-ended questions. Our quantitative results show a modest acceptability of the light logger. A thematic analysis of the open-ended questions reveals that the form factor of the device, in particular, size, weight and stability, and reactions from other people to the wearer of the light logger, were commonly mentioned aspects. In sum, the results indicate the miniaturisation of light loggers and “invisible” integration into extant everyday objects as key areas for future technological development, facilitating the availability of light exposure data for developing personalised intervention strategies in both research, clinical and consumer contexts.

## Introduction

### The importance of light for human health and well-being

Light is a key driver of human physiology and behaviour (1). Exposure to even moderate light levels at night can disrupt our circadian clock, neuroendocrine system and sleep physiology. More specifically, evening and night light can suppress the production of the hormone melatonin, delay the circadian rhythm and affect sleep (2–15). This wide-reaching impact on physiology can lead to significant effects on physical and mental health (16–18). On the other side, daytime light exposure can offset these disruptive effects of light and under some circumstances lead to improvement in alertness.

The non-visual effects of light – in contrast to the vision and visual perception – are mediated by a pathway connecting the retina to the hypothalamus. In addition to the cones and rods, the retina contains the intrinsically photosensitive retinal ganglion cells (ipRGCs), which express the short-wavelength-sensitive photopigment melanopsin (19–25). The ipRGCs differ in how they respond to light to the cones and rods, by preferring wavelengths near 480 nm, prompting the development of novel methods for quantifying light (25).

### Measuring light with wearable devices

Due to the significance of light in controlling fundamental aspects of physiology and behaviour, there has been a drive to develop wearable devices for capturing light exposure in a personalized way, the ‘spectral diet’ (26). To our knowledge, the work by Okudaira et al. (27) was the first to measure light exposure in an ambulatory fashion by mounting a light sensor on the forehead and wrist of an observer. Over the past 40 years, various other ways to capture light exposure have been developed (reviewed in (28)), including pendants, brooches, wrist-worn packages and loggers mounted in the plane of the cornea (e.g., (29,30)).

Light loggers can be used for a variety of purposes (31–34,29,35–49), spanning both research (e.g., associational or intervention studies) and clinical (e.g. compliance to light therapy regime) settings. They differ in their ability to capture quantities related to light accurately (50), with some only capturing the photopic illuminance (lux). As photopic illuminance is related to human brightness perception, but not the non-visual effects of light, the International Commission on Illumination (CIE) standardised the spectral sensitivities for the human photoreceptors, and proposed related units (51). The proposed quantities are called α-opic, where “α” is a placeholder for the L, M and S cones, the rods and the melanopsin-containing ipRGCs in the human retina.

Importantly, different measurement locations may produce different estimates of light exposure which may not be directly relatable or transformable. Furthermore, light loggers worn on the wrist or on the chest may be covered by sleeves or jackets (37,45,48,49), resulting in unreliable data very far removed from actual ocular light exposure, thereby necessitating measurements in the corneal plane.

### Novel light logger capturing α-opic quantities in the corneal plane

More recently, Stampfli et al. (52) developed a novel light logger which attaches to the side of spectacle frames (Figure **1**), placing the sensor in the plane of the cornea, the front surface of the eye. This set-up takes account of a participant’s head movements and replaces manual measurements taken with spectroradiometers at regular intervals in laboratory settings. The *lido* device weights ∼27 g, has a battery life of seven days and captures the α-opic irradiance at 10-sec intervals in an approximately cosine fashion. The *lido* device has been characterised extensively with respect to its metrological properties, including spectral measurements to characterize deviations from α-opic equivalent daylight (D65) illuminance and α-opic irradiance (<10% for various white light sources) and from photopic illuminance (<5% for white light sources), and geometric measurements to characterize the angular response of the device (f2 relative to cosine ∼5%). In addition, the *lido* device was characterized with conjoint measurements with a commercial research-grade device by Ocean Insight, showing good agreement. The *lido* device captures light in the range from approx. 5 lx to 100,000 lx. For further details, the reader is pointed to the publication by Stampfli et al. (52).

**Figure 1:**
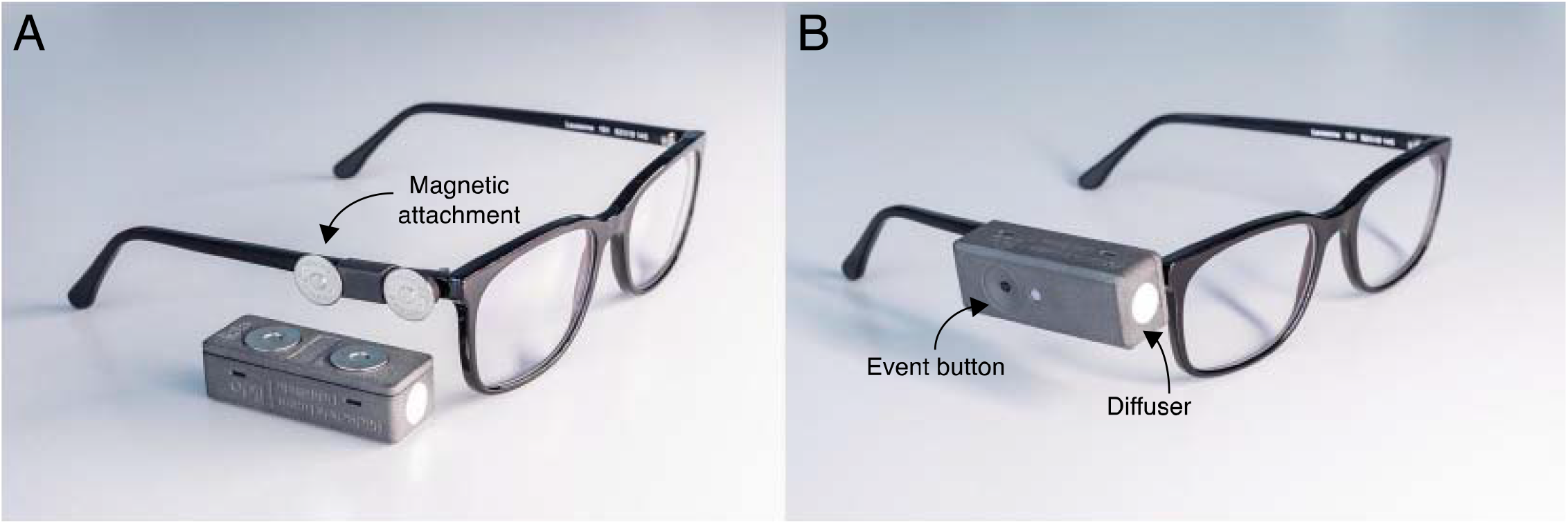
Wearable corneal-plane α-opic light logger. **A** Light logger unattached to spectacle frame, highlighting magnets for secure attachment. **B** Light logger attached to spectacle frames, highlighting diffuser and event button for logging user-defined events. Photos by Licht@hslu.

The *lido* device has the potential to yield biologically-relevant measurements of light exposure in the corneal plane over long periods. Going beyond the technical and analytic validation of the device (53), in this work we examine whether the *lido* device is usable and acceptable in a 24-hour field trial. The goal is to learn whether it can be deployed in large-scale field trials, for which acceptability is a key requirement.

## Materials and methods

### Participants

Participants were recruited from the wider University of Oxford community through an email and internet recruitment campaign targeting participants aged between 18 and 25 years, wearing habitual glasses and living in Oxford. Participants received £25 for their participation in the study.

A total of 18 volunteers (n=18) aged between 18 and 24 years (mean±1SD: 20.1±1.7; 9 female, 9 male; 1 non-binary gender identity) participated in the study. All volunteers were able to read and understand English, were full time students and lived within the Oxford Ring Road. On average, participants reported spending 2.1±1.3 hours daily outside (min: 1, max: 6) on weekdays, and 2.6±1.9 hours outside (min: 0, max: 8) on weekend days (*V* = 50.5, *p* = 0.01503, paired Wilcoxon signed rank test). As commuting to work is a key opportunity for outdoors light, we asked whether participants for their mode of transport bicycle: 8, on foot: 9, mainly work from home: 1).

### Procedure

After completing an initial online screening survey, participants were invited to an in-person visit. Participants’ glasses were fitted with the *lido* by a researcher using a shrinking tube and heat gun. They were given instructions as to how to use the device, including they were told that should they remove the device for a significant amount of time (i.e., longer than 20 seconds), then they should indicate this by pressing the button on the device. They were informed that the glasses emitted a small green flash from an LED light every 10 seconds, this being an indication that the device is working. The participants were instructed to place their glasses on a flat surface when they went to sleep, facing the same direction as they were lying down. This procedure allowed for the capture of light exposure in the sleep environment, limited by the valid range of light levels that can be captured by the *lido* device. After 24 hours the participants were asked to return to the study site to return the devices and complete rating scales and open-ended questions.

### Rating scales and open-ended question

We probed the usability and acceptability of wearing the *lido* device over the 24-hour data collection period using a series of scales. We probed (a) social acceptability of the device using the WEAR (WEarable Acceptability Range) Scale (54), (b) usability using the System Usability Scale (SUS) (55), (c) acceptance using a previously developed scale (56) and (d) subjective experience using the Intrinsic Motivation Inventory (IMI) (57,58), with specific reference to the 24-hour period of data collection (“the task”). For all scales, we used 7-item Likert items capturing agreement (“Strongly disagree”, “Disagree”, “Somewhat disagree”, “Neither agree or disagree”, “Somewhat agree”, “Agree”, “Strongly agree”). In addition to the quantitative scales, we also asked the open-ended question “Do you have any other comments or observations about the device?”. Participants completed the questionnaires via the REDCap system (59,60).

### Analytic strategy

The rating scale data were simply visualised descriptively. The thematic analysis was performed by one author (S.G.) and quality-checked by the senior author (M.S.). We performed thematic analysis using NVivo v12 (QSR International, Burlington, MA), following the steps presented by Braun & Clarke (2006). These steps are as follows: familiarisation with the data; generating initial codes; searching for themes; reviewing themes; defining and naming themes; and producing the report. The *lido* was created by some authors of this paper (development: J.S., B.S.; validation: M.S.). These authors did not not perform any qualitative analysis of the data presented here, which could have led to biases.

## Results

### Subjective ratings

The quantitative data are shown in Figure **2**, showing the rating data for the WEAR (WEarable Acceptability Range) Scale, the System Usability Scale (SUS), acceptance using a previously developed scale and subjective experience using the Intrinsic Motivation Inventory (IMI). As the scales probes a large number of items, we highlight only the most salient aspects here, concerning comfort and public perception.

**Figure 2:**
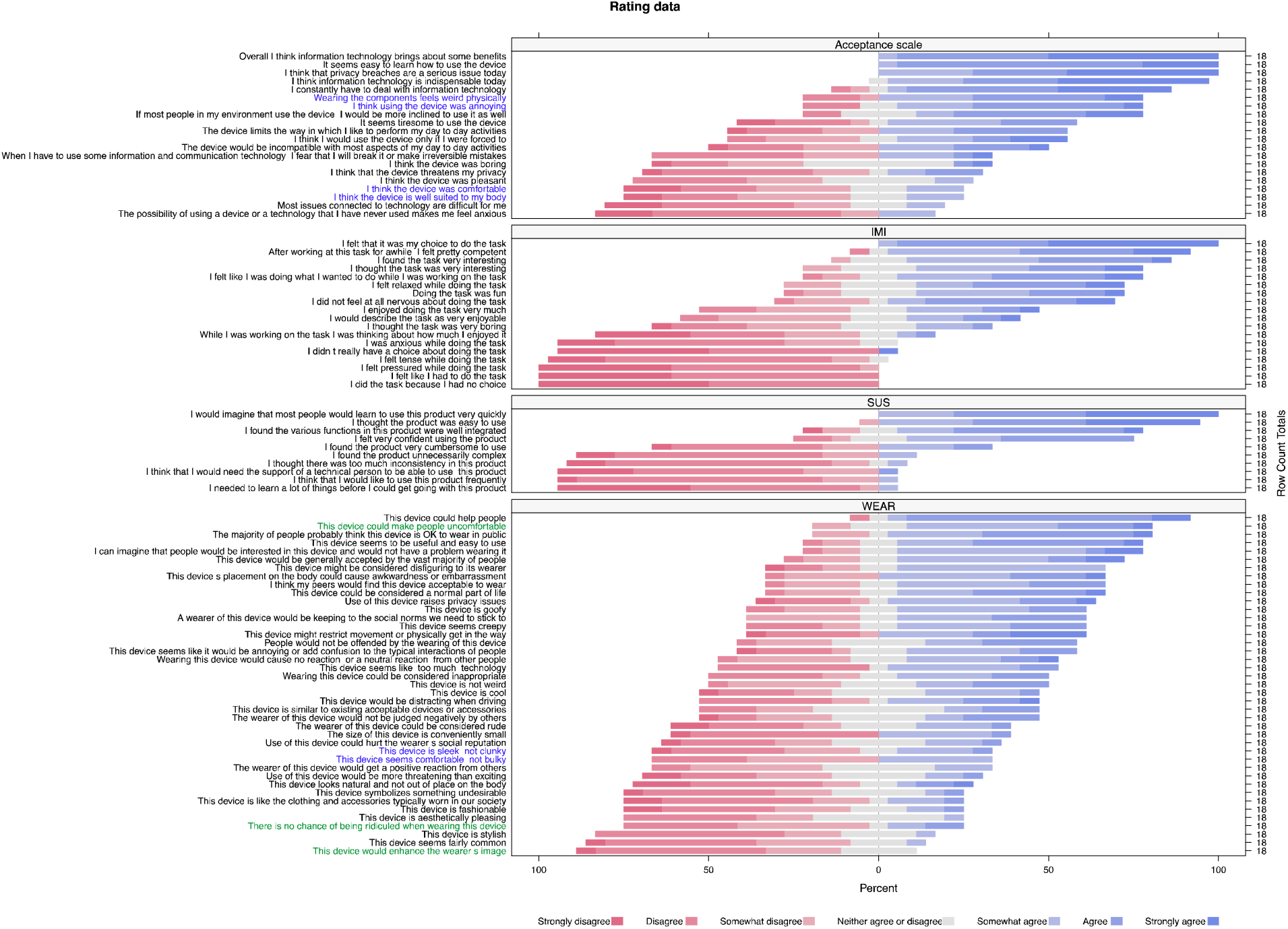
Rating data. Highlight colours correspond to specific areas highlighted in the text (blue: convenience and comfort of using the device, green: perception of others)

### Convenience and comfort of using the light logger

The light logger was found to be annoying, uncomfortable, aesthetically not pleasing and not fashionable, and bulky. We highlight representative items in Figure 2 in colour: “Wearing the components feels weird physically” (high agreement), “I think using the device was annoying” (high agreement”, “I think the device was comfortable” (low agreement), “I think the device is well suited to my body” (low agreement), “This device is sleek, not clunky” (low agreement), and “This device seems comfortable, not bulky” (low agreement).

### Perception of light logger users

Participants rated the perception that users, scoring high agreement on the item “This device could make people uncomfortable” and low agreement on the items “There is no chance of being ridiculed when wearing this device” and “This device would enhance the wearer’s image” (Figure **2**).

### Qualitative results

Eleven (n=11) of the 18 participants completed the open-ended feedback. The thematic analysis revealed three main clusters: (1) size and weight of the device, (2) reactions of others, (3) positive feedback. The themes and sub-themes contained within these are presented in Table **1**, along with illustrative quotes, and are detailed below.

**Table 1.** Themes and sub-themes identified from thematic analysis

| ***Theme*** | ***Sub-themes*** | ***Illustrative Quotes*** |
| --- | --- | --- |
| Size and weight of the device | Instability on the glasses frame | “In my dance class I was unable to participate in all the steps as a lot of jumping is involved and I feared the device would fall off.” |
|  | Affecting the tilt of the glasses | “The weight is noticeable, as it causes a slight imbalance of the glasses.” |
|  | The dimensions of the device being too great | “Which made it very cumbersome to wear.” |
|  | Suggestions for improvements | “An idea would be to have a weight on the other side of the specs as well so that it remains balanced.” |
| Reactions of peers | Positive reactions | “All my friend asked me what that was but they took it in a fun way and actually enjoyed the device and knowing more about it.” |
|  | Negative reactions | “My tutors feared that it was a secret recording device with a camera so they were very suspicious in the start.” |
| Positive feedback | Positive feedback regarding physical aspects of the device | “The device was mostly comfortable.” |
|  | Usefulness of the information recorded by the device | “I think the information it can record is very useful.” |

Size and weight of the device

#### Instability on the glasses frame

One criticism of the prototype device was that it felt unstable on the participants’ glasses, with one participant “[fearing] the device would fall off” during a dance class, and another stating the device felt like it was going to fall off even when they were working (“made it want to fall off whenever I was working [and] turning my head”). This suggests that for both vigorous (dancing) and non-vigorous (working) activities, the device does not feel secure when attached to a glasses frame.

#### Affecting the tilt of the glasses

A key criticism seen in replies to the open-ended question are those referencing the way in which the device causes glasses frames to tilt. 8/11 participants who completed this question referenced this issue. This device was cited as having “tilted” a participant’s glasses which “affected [their] vision a little bit”. The weight of the glasses, and the resulting tilt of some participants’ glasses also appeared to have a negative effect on concentration, with one subject stating: “Because the device is heavy I could feel my specs leaning on one side which made it difficult to focus on studies, because you are wearing diagonal specs”.

#### The dimensions of the device being too great

The idea of the device being too “bulky” was also a theme that reoccurred throughout participant’s feedback. This was not referencing the effect of the size and weight of the device on the participants’ glasses, but rather on the participants themselves. Participants could have been “aware of the device for most of the task” due to its size. One participant called the device “[v]ery cumbersome to wear”.

#### Suggestions for improvements

Of those participants who raised an issue with the tilt of their glasses caused by the device, a number also offered ways they thought this could be overcome. All these suggestions involved adding a “load on the other side” of the glasses frame being used, to offset the weight of the device, and keep the frames “balanced” (“[t]his can be solved by adding a counterweight on the opposite arm of the glasses”). To address the size of the device being too great, a suggestion was given to move the position of the device such that participants would no longer “glance at the device”. This would detract from its distractions during activities like “cycling” (“could potentially be moved further back so it can’t be seen”).

### Reactions of others

#### Positive reactions from peers

A few participants described others’ positive reactions to the device. These appear to centre on curiosity in the device, with friends interested in “knowing more about it”: “She was quite intrigued knowing I was wearing a [dosimeter] as part of an experiment – neutral to positive reaction”.

#### Negative reaction from others

One theme that was seen was a suspicion from others as to the purpose of the device. This was seen both for peers and members of authority. Although the device does not record anything other than light and tilt measures, those who are not participating in the study may not be aware of this. This may lead to another issue, that regarding negative attention such as “having people stare at the device while you wear it” which may act to make participants “feel a little uncomfortable”. This is illustrated in the following quote: “The placement of the device and the small circle on the front meant that some people assumed that it was a video recording device, which made them feel uncomfortable due to privacy issues”

### Positive Feedback

#### Positive feedback regarding physical aspects of the device

There were also comments relating to advantages of the device itself and its usage. One participant cited it as being “mostly comfortable”. Another stated that it was “unobtrusive and easy to use”. This latter participant had thicker glasses frames and this may have influenced why they thought it was more subtle than others with smaller/thinner frames.

#### Usefulness of the information recorded by the device

Another theme which was seen was reference to the use of the information collected by the device. The device was seen as “very useful” by some participants. One avenue which was referenced as being particularly useful was to track “all of the artificial light from electronic devices” and suggests that participants may be interested in this device for more clinical reasons. One participant wrote: “Might be very beneficial for […] eye health, sleep circadian rhythms, and brain health! I think the device therefore has a lot of potential in its future uses!”

### Objective light measurements

Throughout the trial, we collected actual corneal light exposure data. We present the quantitative data collected for 17 participants in Figures 3 and 4. One participant (P03) was excluded from this analysis due to abnormal and physiologically implausible patterns of light exposure. The exemplary 24-hour time courses of light exposure are shown in Figure 3, with each panel corresponding to one participant. The data was thresholded to remove melanopic equivalent daylight illuminance (mEDI) values below 1 lux, in accordance with the estimated valid and trustworthy range of light levels. This enabled the identification of a main sleep or “lights off” period for each participant, which was scored visually by one author (C.G.). An overview of variations in melanopic light exposure across the 24-hour period for all participants can be observed in Figure 4, where a 1-hour centered moving average was applied to the data for each participant for smoothing. As shown, mEDI levels progressively drop towards individuals’ bed time, to increase again after wake. Figure 4 also illustrates the relationship between mEDI and photopic illuminance, calculated for day periods.

**Figure 3:**
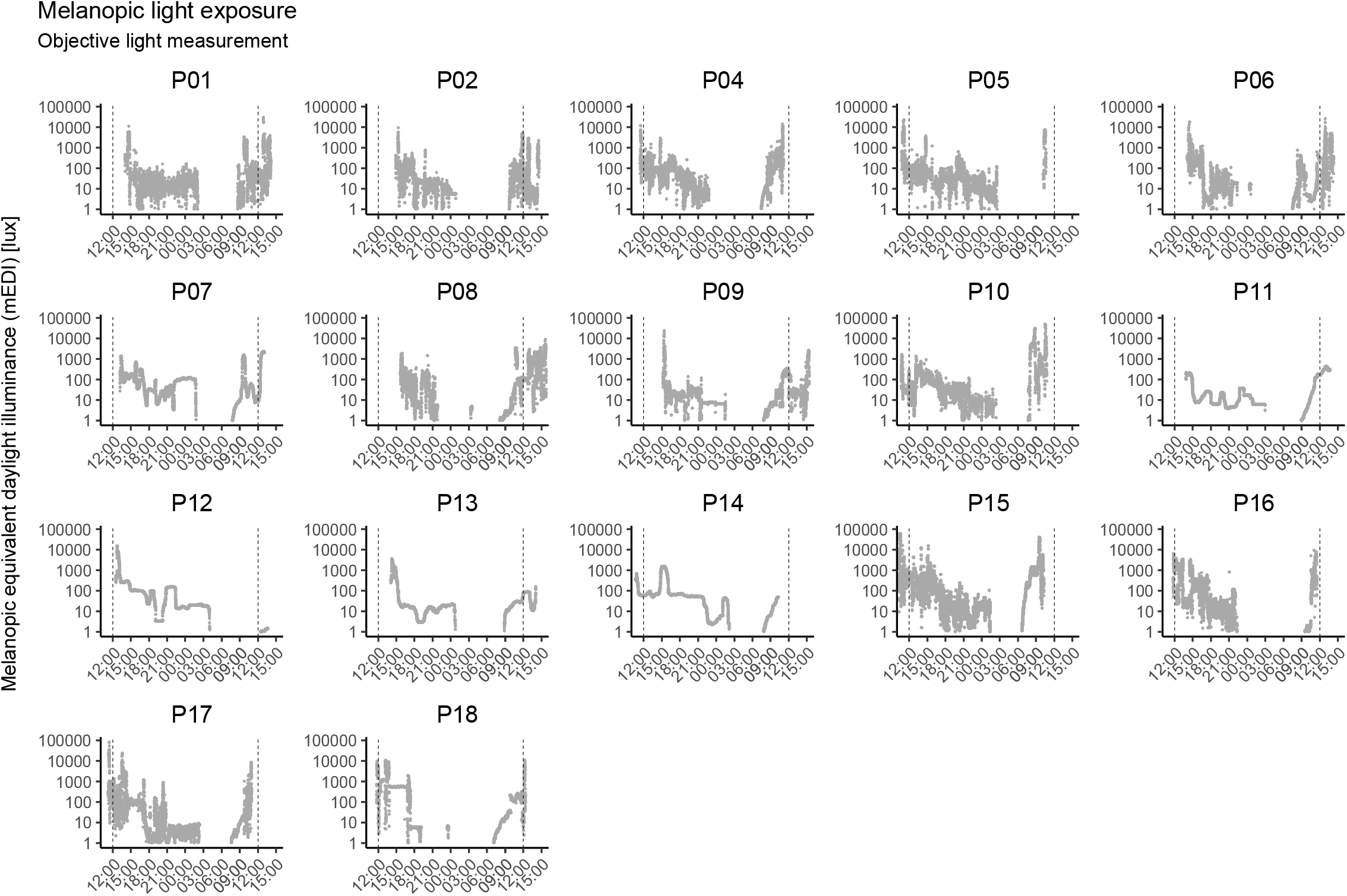
Objective light exposure data for participants with valid data (n=17). Yellow regions show visually scored main sleeping/”lights off” periods.

**Figure 4:**
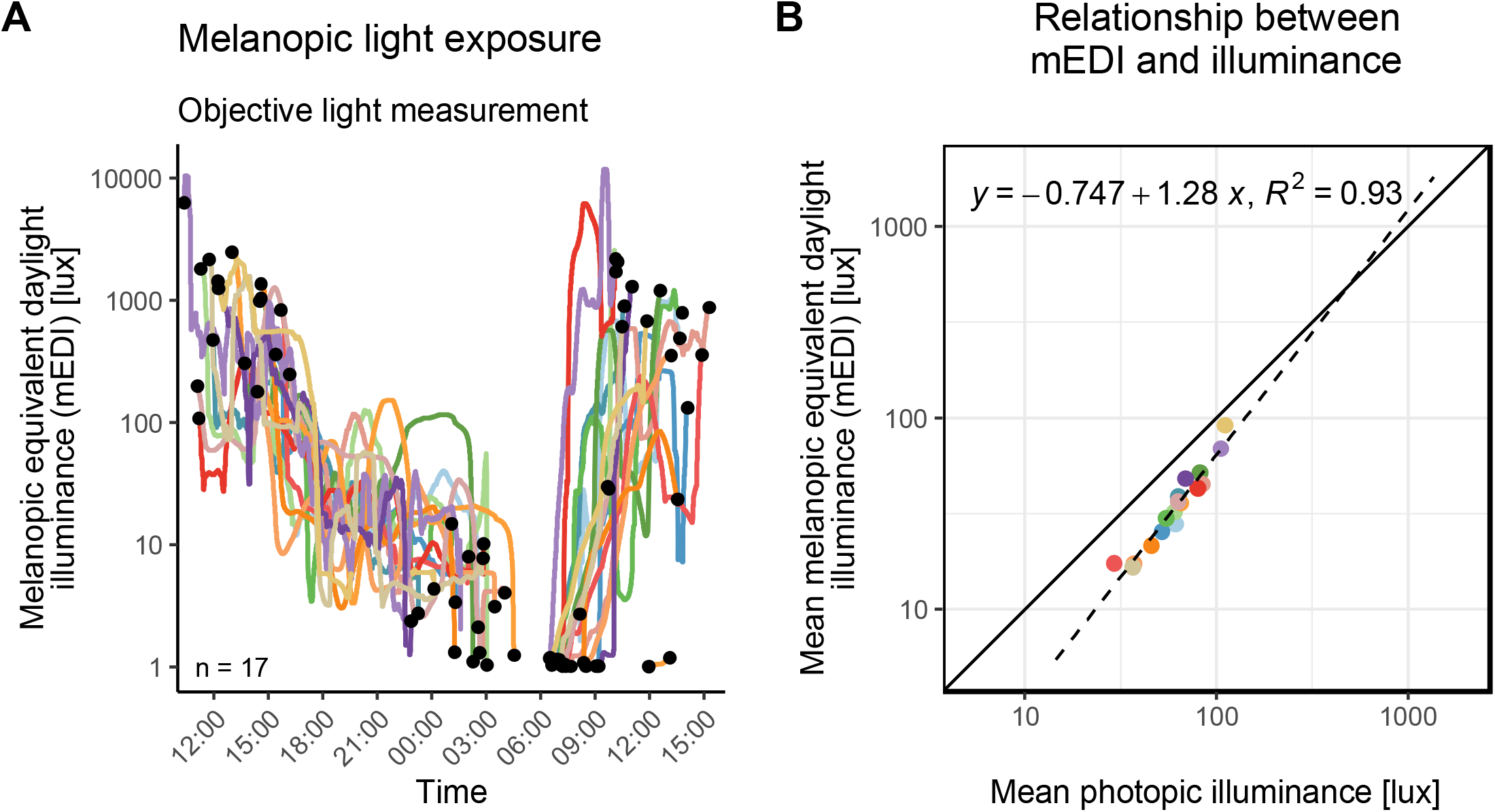
Melanopic light exposure across the 24-hour period and during day periods. (A) Changes in melanopic equivalent daylight illuminance (mEDI) across 24 hours (n=17). Individual lines represent 1-h centred moving averages and black dots indicate start and end of data collection and sleep periods. (B) Correlation between mean photopic illuminance and mean equivalent daylight illuminance (n=17) participants. Individual points represent mean values calculated for “day” periods.

## Discussion

### Known limitations

A key limitation of this study is the duration of the data collection and wearing period, which was restricted to 24-hours. While a 24-hour observation duration may be useful for obtaining exemplary data for a participant, it is expected that there are significant inter-daily variations in light exposure. As a consequence, our quantitative and qualitative data may provide only a partial insight into the usability and acceptability with longer data collection periods. In the assessment of sleep-wake rhythms using actigraphy, for example, data collection periods of >1 week are recommended (62–64). Given that there are differences in light exposure between weekdays and weekends (65–69), it is expected that a similar or longer measurement duration would be necessary to yield useful light logging data. Additionally, metrics to summarise light exposure, such as the recently proposed Light Regularity Index (70), require longer data collection periods. It is expected that if anything, the concerns raised by the participants during a 24-hour trial will be similar, if not worse, in longer trials.

Although providing us with important information about the participants’ opinion regarding the device, a major limitation is the sample size for the qualitative analysis. Only eleven of the eighteen participants provided this type of data. This meant that searching for consistent themes across participants was challenging. For some of the themes (positive reactions from peers, instability on the glasses frame, positive feedback regarding physical aspects of the device, and usefulness of the information recorded by the device) there were only two examples available. Although this gives us insight into the opinions of some participants, a larger investigation may be necessary to uncover these aspects in detail. To maximise the completeness of the qualitative data, it may be useful to require participants to answer the open-ended question included in the surveys.

The approach taken here focused on the use a single light logger form factor, that of the *lido* device worn as an attachment to spectacle frames. As a consequence, the data presented here are not indicative of the “absolute” acceptability and usability of this specific instance of the device. Instead, the study primarily provides qualitative insights into which design features may be targets for improvement. To advance our understanding of the practical needs of light logging forward, it may be advisable to investigate other form factors, e.g., brooches or pendants, in a systematic investigation, Such an investigation, which does not necessarily require the availability of functional devices but could work with mock devices, could deploy focus-group methodology to discover potentially novel ways of designing a wearable light logger. Importantly, we believe that the choice of light logger for a given research or clinical application follows a trade-off of form factor, usability, long-term deployability and demands on data quality, fidelity and physiological relevance.

Finally, the sample in this study consisted of 18 young, healthy and cooperative students in Oxford, UK, representing a “WEIRD” (White, Educated, Industrialized, Rich, and Democratic) sample (71). There may be idiosyncrasies in the subjective evaluation of usability and acceptability in of the corneal-plane devices used here due to the moderate sample size and limited geographical context. The results of this study indicate modest acceptability of the devices, which we expect may decrease further in clinical populations, or other samples. Consequently, the usability and acceptability of light loggers – of any form factor – may require further investigation across diverse samples and diverse contexts.

### Towards convenient, continuous all-weather personal light logging

The thematic analysis undertaken produced three themes: size and weight of the device; reactions of others; and positive feedback. The majority of feedback (8/11) included negative comments regarding the weight of device. Not only did this include concerns about the size of the device and the consequences of this, but also suggestions on how to correct its instability and the tilt of glasses frames. These combined suggest that the device is currently too large and heavy. This feedback should be integrated by researchers and developers when developing new wearables, as it emphasizes the importance of size and weight both from a practical and acceptable standpoint.

The device has an IP20 rating, which means that it is product and will be resistant to objects >12 mm in size. However, this also means that it has no protection against liquids and will be susceptible to damage if it comes into contact sprays of water. This meant that we could not perform data collection on days when it was raining heavily. However, it is likely that participants would have spent a greater amount of time indoors anyway on days with particular rain. It is more likely to have been affected on days in which rain was light, as individuals may have performed normal daily activities, including outdoor ones, but the device would still need to be removed from the glasses frame in this instance. In this situation, the light data gathered would not reflect the “spectral diet” of the affected individual. Future devices should consider weather-proofing the devices comprehensively to allow for continuous all-weather light logging. The *lido* device is mounted on spectacle frames, limiting the range of use cases, as it requires the wearer to habitually and continually wear glasses. Participants were instructed to place their glasses on a flat surface when they went to sleep, thereby allowing us to collect data in the sleep-environment. Due to the lower end of the dynamic range of the *lido* device (∼5 lx), this procedure represents a compromise in being able sample light in the sleep environment.

There have been recent advances in towards miniaturization of light sensors with spectral capabilities (73–77). These may result in new opportunities for developing light loggers and dosimeters that are nearly invisible or at least to some degree integrated into common objects (e.g., earrings, spectacles). Such developments could aid in addressing some of the challenges brought forward by the participants here, specifically regarding size and weight, and intrusiveness, and also mitigate the potential loss of data due to sensors being covered by sleeves or other clothing.

## Conclusion

Wearable light loggers represent a methodology to capture light exposure in a personalised fashion. Here, we examined the acceptability of a spectacle-attached light logger capturing α-opic irradiance. Through quantitative and qualitative analysis, we found modest acceptability in real-world conditions. The qualitative analysis highlights size and weight as key targets for improvement, providing imperatives for developing novel light sensors with a smaller footprint to enable the “invisible” capture of visible optical radiation in everyday settings.

## Supporting information

Plain Language Summary

## Data Availability

All data, code and materials are available on GitHub (https://github.com/TUMChronobiology/lido-acceptability/).

## Statements

Statement of Ethics

The research project was reviewed and approved by the Medical Sciences Interdivisional Research Ethics Committee (MS IDREC) of the University of Oxford (R79098/RE001, R79102/RE001). All participants gave written informed consent. The research was conducted in accordance with the World Medical Association Declaration of Helsinki.

## Author Contributions

*Conceptualization*: Manuel Spitschan

*Data curation*: Manuel Spitschan

*Formal Analysis*: n/a

*Funding acquisition*: Manuel Spitschan

*Investigation*: Eljoh Balajadia, Sophie Garcia, Carolina Guidolin, Manuel Spitschan

*Methodology*: Eljoh Balajadia, Sophie Garcia, Manuel Spitschan

*Project administration*: n/a

*Resources*: Janine Stampfli, Björn Schrader

*Software*: Janine Stampfli, Björn Schrader, Carolina Guidolin,

*Supervision*: Manuel Spitschan

*Validation*: n/a

*Visualization*: Carolina Guidolin, Manuel Spitschan

*Writing – original draft*: Manuel Spitschan

*Writing – review & editing*: Eljoh Balajadia, Sophie Garcia, Janine Stampfli, Björn Schrader, Carolina Guidolin, Manuel Spitschan

## Data Availability Statement

All data, code and materials are available on GitHub (https://github.com/TUMChronobiology/lidoacceptability/).

