## Supplementary material for "Usability and acceptability of a corneal-plane α-opic light logger in a 24-hour field trial": Plain Language Summary

This study looks at the effects of light exposure on people's biology and behaviour. It is well-known that exposure to light helps our bodies know when to sleep and wake up, and controls the production of a hormone called melatonin. While previous studies have been done in controlled laboratory settings, researchers have developed new ways to study the effects of light in more natural settings.

To do this, researchers created a wearable device that attaches to the frames of glasses and measures the type of light that enters the eye. They wanted to see if this device was usable and acceptable to wear for a full day in real-world conditions.

The study involved 18 university students in the UK, who wore the device for 24 hours. Researchers asked them questions about how easy it was to wear, how comfortable it was, and how they felt about wearing it in public.

The results showed that while most students found the device acceptable to wear, there were some concerns about its size, weight and stability. Some students also reported feeling self-conscious about wearing the device in public.

Overall, the study suggests that wearable devices like this one could be useful for studying the effects of light exposure in real-world settings. However, researchers will need to address concerns about the device's design and how it is perceived by others in order to make it more user-friendly and widely adopted.
